## Supplementary Figures for "Pin-pointing the key hubs in the IFN-γ pathway responding to SARS-CoV-2 infection"

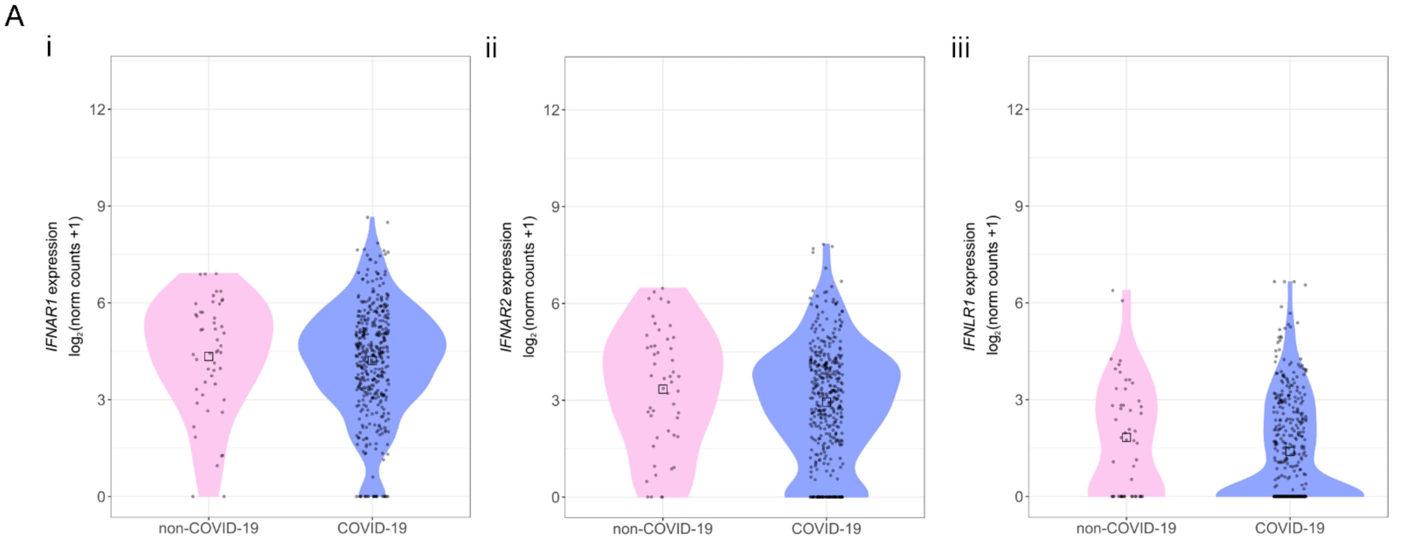


**SUPPLEMENTARY FIGURE S1. Expression of genes encoding for the receptors involved in IFN-I and IFN-III signaling pathways in non-COVID-19 and COVID-19 patients.** Gene expression analysis (log2 (norm counts +1)) for IFNAR1 (i), IFNAR2 (ii), and IFNLR1 (iii) in COVID-19 (purple) vs. non-COVID-19 (pink) patients from the GSE152075 dataset, assessed by RNA-seq. p-values correspond to Wilcoxon rank-sum test. Black squares represent the median.

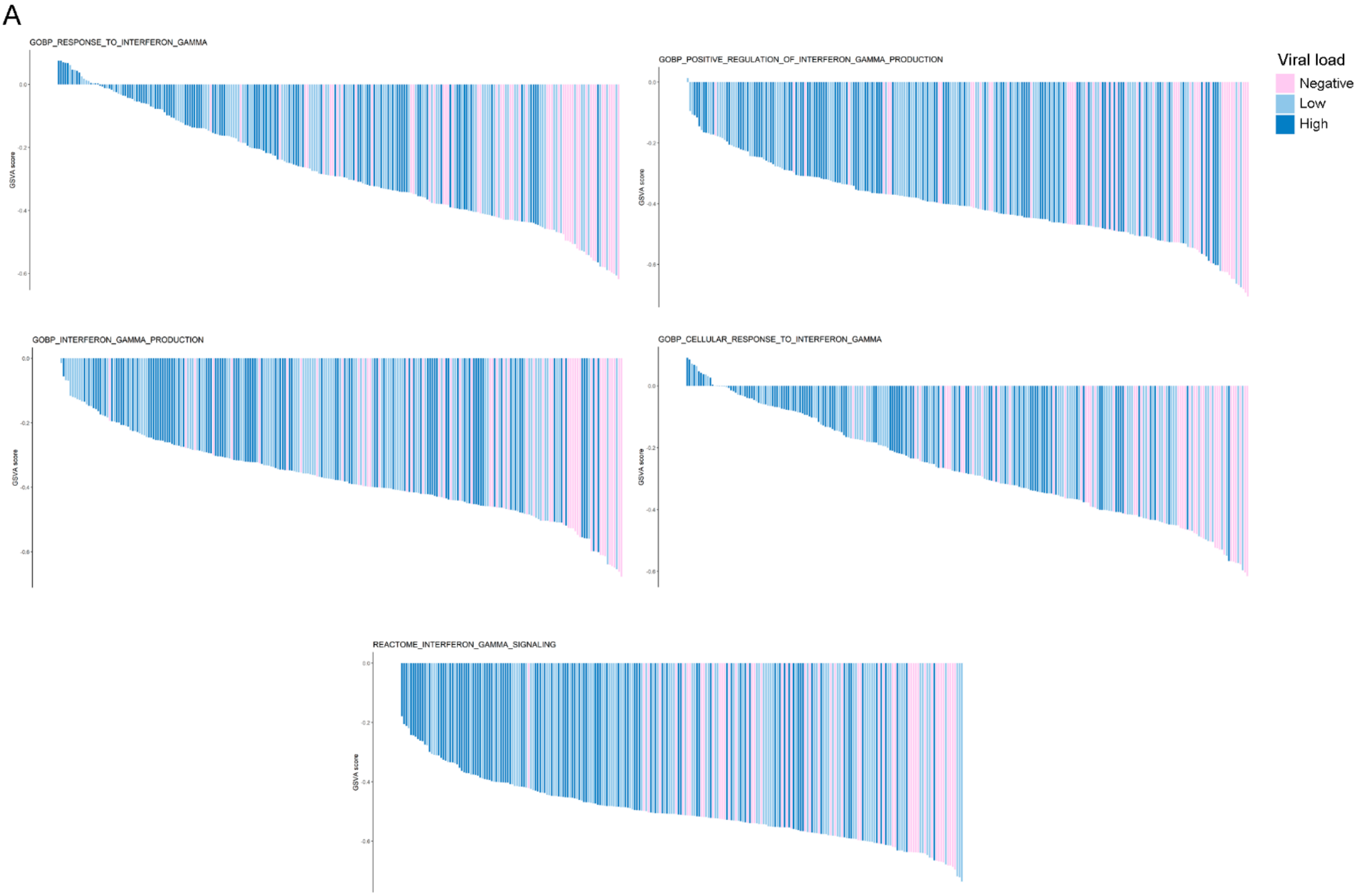


**SUPPLEMENTARY FIGURE S2. Global assessment at the transcriptional level of pathways related to IFN-γ production, signaling and regulation of response in COVID-19 positive and negative patients.** Waterfall plots of selected genesets that were activated in COVID-19 patients vs. non-COVID-19 patients. Patients are ordered from the highest to the lowest GSVA score in each gene-set.

SUPPLEMENTARY FIGURE S3

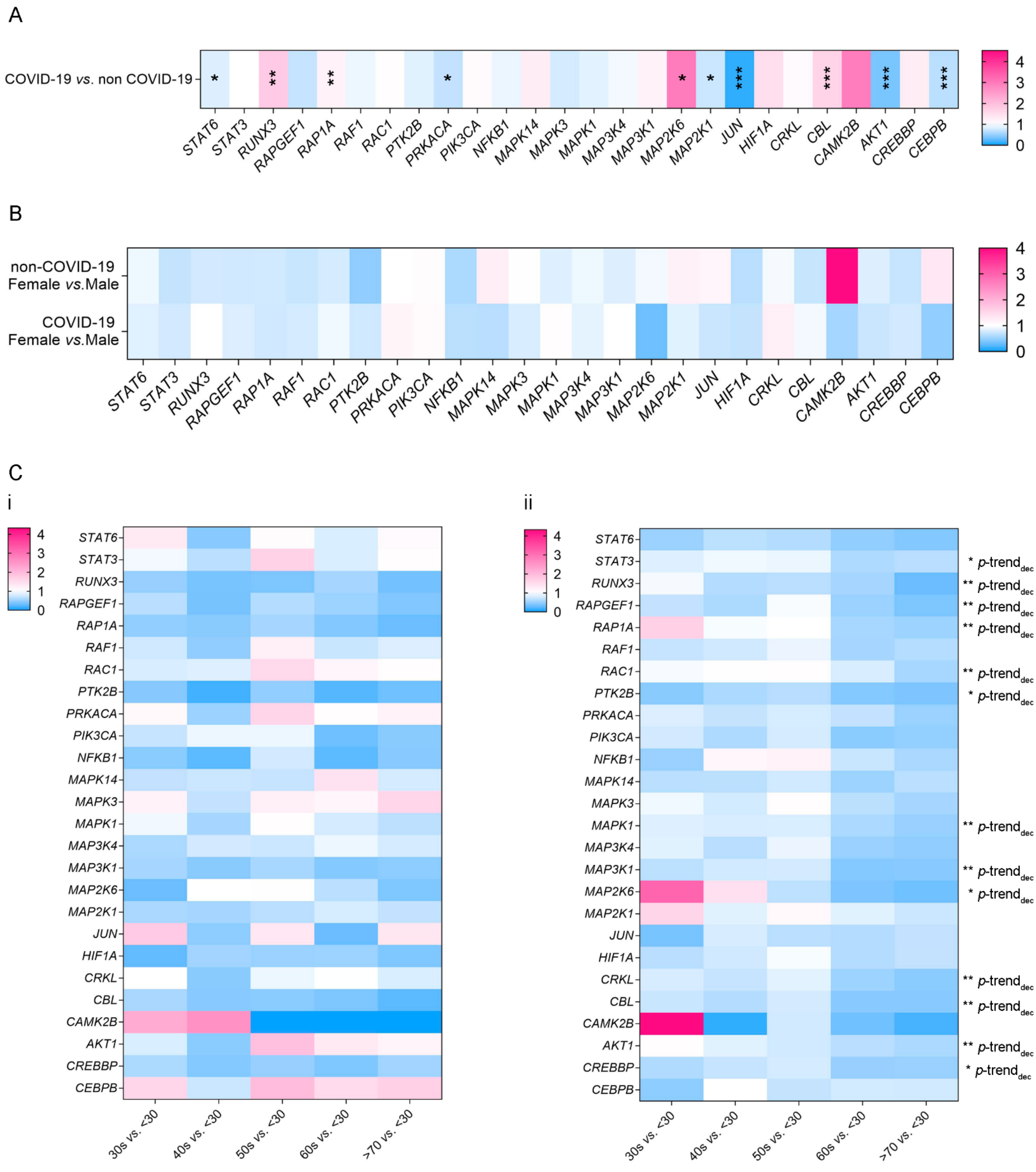


**SUPPLEMENTARY FIGURE S3. Expression of genes belonging to the non-canonical IFN-γ pathway in non-COVID-19 and COVID-19 patients from the GSE152075 dataset.** Heatmaps depicting the fold change (high = pink; low = blue) for gene expression of genes belonging to the non-canonical IFN-γ pathway between COVID-19 vs. non-COVID-19 patients (A); and female vs. male (B), and age groups 30s, 40s, 50s, 60s & 70s vs. <30 (C) in non-COVID-19 (i) and COVID-19 (ii) patients, assessed by RNA-seq. For (A) & (B) p-values correspond to Wilcoxon rank-sum test, for (C) p-values correspond to decreasing Jonckheere-Terpstra trend test. Statistical significance *p < 0.05; *p < 0.01; ***p < 0.001.

SUPPLEMENTARY FIGURE S4

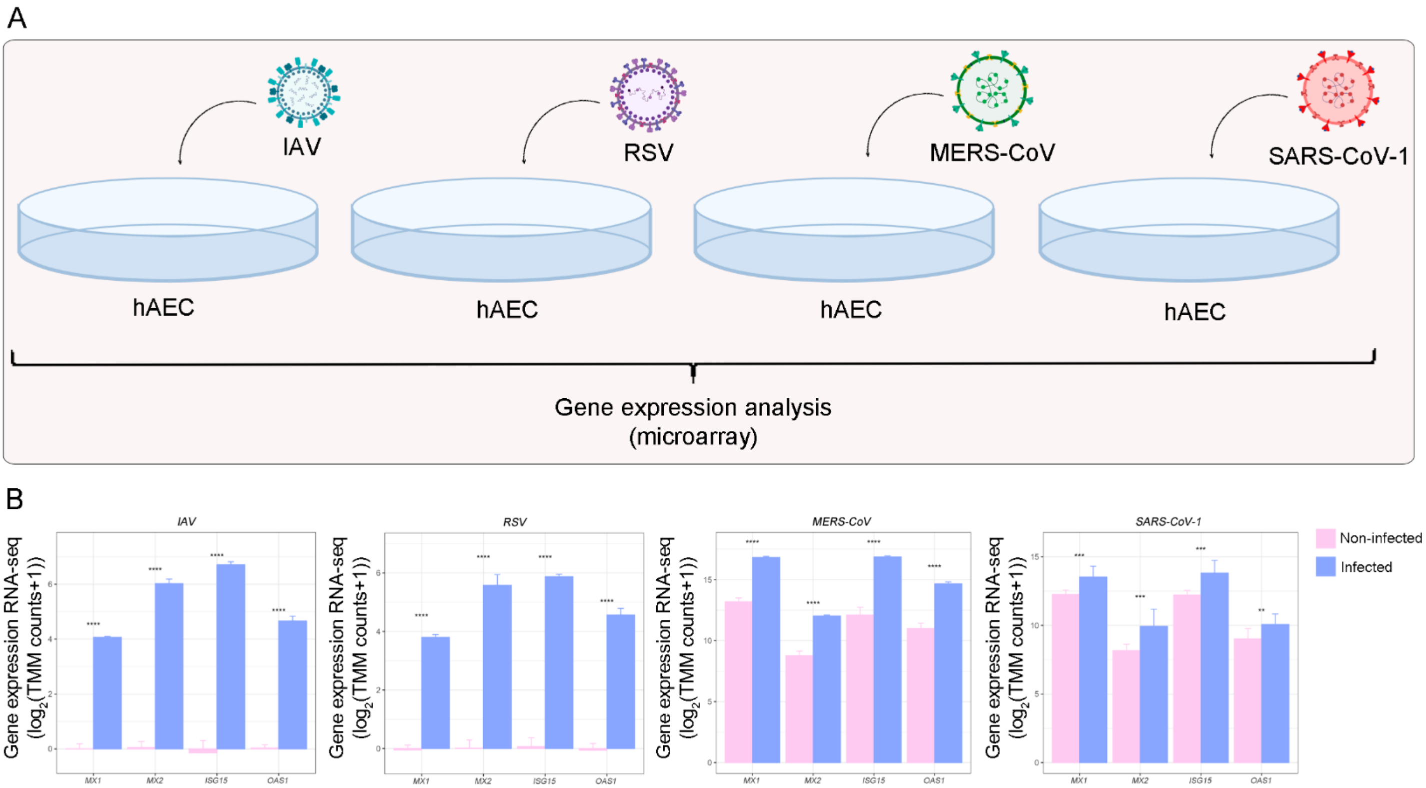


**SUPPLEMENTARY FIGURE S4. MX1, MX2, ISG15 and OAS1 expressions in human primary airway epithelial cells (hAEC) that were infected with influenza A (IAV) (2×105 PFU, 24 h), respiratory syncytial virus (RSV) (1×106 PFU, 48 h), Middle East respiratory Syndrome (MERS-CoV) (MOI 5, 48 h) or SARS-CoV-1 (MOI 2, 48 h).** A) Schematic representation of the experimental design. B) MX1, MX2, ISG15 and OAS1 expressions in infected (purple) vs. mock-treated (pink) hAEC human cell lines, assessed by microarrays. Data analyzed for IAV and RSV were obtained from the GSE32138 dataset (n = 8). Data analyzed for SARS-CoV-1 and MERS-CoV were obtained from the GSE47963 (n = 20) and GSE100504 (n = 10) datasets, respectively. Student's t test was performed to determine statistical differences. Statistical significance $*p < 0.05$; $**p < 0.01$; $***p < 0.001$.
